# Analysis of communities of countries with similar dynamics of the COVID-19 pandemic evolution

**DOI:** 10.1101/2021.01.18.21250050

**Authors:** Emiliano Alvarez, Juan Gabriel Brida, Erick Limas, Lucia Rosich

## Abstract

This work addresses the spread of the coronavirus through a non-parametric approach, with the aim of identifying communities of countries based on how similar their evolution of the disease is. The analysis focuses on the number of daily new COVID-19 cases per ten thousand people during a period covering at least 250 days after the confirmation of the tenth case. Dynamic analysis is performed by constructing Minimal Spanning Trees (MST) and identifying groups of similarity in contagions evolution in 95 time windows of a 150-day amplitude that moves one day at a time. The number of times countries belonged to a similar performance group in constructed time windows was the intensity measure considered. Groups’ composition is not stable, indicating that the COVID-19 evolution needs to be treated as a dynamic problem in the context of complex systems. Three communities were identified by applying the Louvain algorithm. Identified communities analysis according to each country’s socioeconomic characteristics and variables related to the disease sheds light on whether there is any suggested course of action. Even when strong testing and tracing cases policies may be related with a more stable dynamic of the disease, results indicate that communities are conformed by countries with diverse characteristics. The best option to counteract the harmful effects of a pandemic may be having strong health systems in place,with contingent capacity to deal with unforeseen events and available resources capable of a rapid expansion of its capacity.

## 1. Introduction

The COVID-19 pandemic continues to batter the world, with more than 70 million positive cases and almost two million deaths by the end of December 2020 (see Santiago et al., 2020). This study extends Alvarez et al. (2020), in which the authors analyze the evolution of the COVID-19 in a dataset of 191 countries. The paper utilizes a clustering method based on the correlation distance and a Minimal Spanning Tree (MST), from which they obtain a Hierarchical Tree (HT). The algorithm identifies three clusters, each one associated with a different shape in the infections curve. The authors suggest that the COVID-19 propagation is a complex phenomenon whose dynamics can be better understood within the framework of the complex systems theory.

Besides the already-mentioned Alvarez et al. (2020), other authors have also employed non-parametric techniques in order to analyze the COVID-19 dynamics. Zarikas et al. (2020) analyze the set of 30 countries with the highest number of COVID-19 cases and identify four main clusters during the period between the 22nd of January 2020 and the 4th of April 2020. One of the groups is characterized by an abrupt increase in the number of active cases. A second cluster is associated with both an increase and a flattening in the curve of infections. A third group clustered the countries with the lowest number of cases and finally, the fourth group showed an abrupt increase during the last part of the period analyzed.

A K-means clustering study is presented in Chandu (2020), in which the author analyses countries with at least 1,000 COVID-19 cases (for a discussion on the clustering algorithm see Fahim (2020)). The algorithm grouped the set of countries into two clusters. It was observed that one of the clusters, integrated mainly by European countries, Australia, USA and Canada was characterized by a high fatality rate.

On the same line, Machado and Lopes (2020) analyze a data set of 79 countries following two approaches, heuristic models and hierarchical clustering. The authors point out that the emergence and propagation of COVID-19 is an example of a complex system. A consequence of this observation is that the analysis of the new Coronavirus disease requires a methodological framework that assumes the fact that the World is currently facing a rare and extreme event. As we will explain in the methodological section, in the first stage of this work we analyze the spread of the coronavirus as a complex network in which each link represents how similar the coronavirus dynamics is between any pair of countries. Additionally, network analysis is applied in order to identify the intensity of the similarity among communities.

The above-mentioned studies were carried out for different time frames and countries obtaining diverse results. This suggests that findings related to the evolution of the pandemic are sensitive to these factors. Considering that countries presented a greater variety of trajectories as the disease progressed, this study is based on analyzing the evolution of contagions from a dynamic perspective.

The remainder of this paper is organized as follows. Section 2 introduces data, Section 3 presents the methodology, whereas section 4 presents the empirical results. Finally, section 5 concludes.

## 2. Data

This study is based on the COVID-19 data of active cases per population published by “Our World in Data” (Roser et al., 2020). This source is put together with information from a variety of sources: European Centre for Disease Prevention and Control, government reports, Oxford COVID-19, Government Response Tracker, World Bank - World Development Indicators, United Nations Statistics Division and Eurostat.

In particular, our main variable of interest is “new cases per million”, reported by the European Centre for Disease Prevention and Control. We compare the evolution of the disease in countries that by December 2 had data for more than 250 days after the confirmation of the tenth case. With the aim of having a balanced panel that allows to perform distance metrics, all countries’ data of new cases per million are considered for the same time span. This implies that each country’s COVID-19 cases are considered in a different period. For example, for China, the first country that identified the disease, the period between January 22 and September 27 2020 is considered. The country with the most recent data is Mali, which by March 27 had more than 10 confirmed cases, being analyzed until December 1st.

A seven-day moving average smoothing centered on the day of reference was made to the original data, reducing the time series to the central 244 data. Additionally, some countries were removed due to having records of negative new cases after the applied smoothing (Uganda, San Marino, Mauritius, Monaco, Lithuania, Luxemburg, Jordan and Ecuador). Moreover, Tanzania was removed due to her stopping cases report since May. Consequently, the analysis is based on 124 series of 244 observations each.

Additional information from “Our World in Data” was considered for groups’ analysis. In particular, socioeconomic indicators-Gross Domestic Product (GDP) and Human Development Index (HDI)-, demographic/population density, median age, life expectancy – and others related to the pandemic – Government response, testing policy, contact tracing, etc.-were considered (see Appendix 1).

## 3. Methodology

Following the methodology developed by Mantegna (1999), in this study the coronavirus propagation is formulated as a network problem, where each country would be represented as a node, and the relationship between each pair of countries as a link.

### 3.1. Time windows networks

On a first stage, distances between the time series of new cases per million of inhabitants are calculated to construct complete adjacency matrices. Pearson’s correlation is the select measure of distance, which summarizes the grade of similarity of new registered cases per million of inhabitants between countries at each considered time window. Given that Pearson’s correlation is an invariant to scale measure (Aghabozorgi et al., 2015), countries that had similar shapes at their trajectories of propagation but differ in the proportion of the affected population will be considered similar and likely to cluster. Following Mantegna and Stanley (2000), Pearson’s correlations between the n x n pairs of chosen countries is computed (see Equation 1) as follows

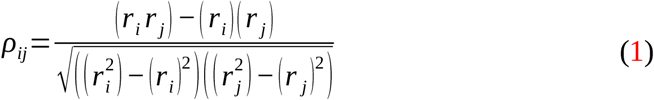

where *r*_*i*_ is the number of new daily cases in country *i* and (*r*_*i*_) is the average value of *r*_*i*_ during the considered period. Then, the correlation matrix is built with the correlation coefficients *ρ*_*ij*_. By definition *ρ*_*ij*_ takes values in the interval(*−*1,1), where *−*1 means completely anti-correlation, 1 complete correlation and 0 that the two variables are uncorrelated. This matrix is symmetrical, with *ρ*_*ij*_=1 in this main diagonal. As it is well known, the Pearson correlation coefficient (1) does not fulfill the three axioms that define a Euclidean metric. For this reason, the correlation matrix is transformed into the correlation distance matrix according to the Equation 2

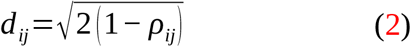

which fulfills the three axioms of an Euclidean distance:

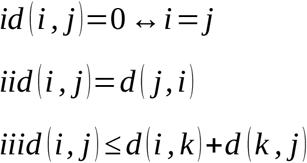

Subsequently, Prim’s algorithm (Prim, 1957) is applied to adjacency matrix to obtain Minimal Spanning Trees (MST). Being introduced to graph theory by Kruskal (1956) and Prim (1957), MST have been a widely used tool (Limas, 2019; Górski et al., 2008; Kwapień et al., 2009; Rešovský et al., 2013; Wang et al., 2013), mainly because it simplifies network analysis by selecting the most relevant bounds. Indeed, MST are characterized for representing the core information of a complete network with n nodes by selecting the n-1 links that minimize the overall distance.

Prim’s algorithm establishes a procedure in successive stages for the selection of MST links. Taking the information from a complete adjacency matrix, at each step a node is selected and incorporated to the network. The criteria is to choose, from the not connected nodes, the one that has the shortest distance to a connected one. At the end of the process all nodes (n) are connected by n-1 links in a network that has the smallest possible total length (Prim, 1957).

Subsequently, the single linkage method is applied to obtain a subordinant ultrametric distance matrix from constructed MST. This graph method is a particular agglomerative hierarchical clustering algorithm. It starts by considering all the nodes of the network as subgroups. In successive stages, the less distant subgroups are joined, the distance between the new subgroup and the rest is determined based on the nearest neighbor criteria. Additionally, every subordinate ultrametric distance matrix can be represented by a Hierarchical Tree (HT) or dendrogram (Gan et al., 2007).Finally, the pseudo T^2^ and CH cutting criteria are considered to determine the optimal number of groups, the highest number of suggested groups with a maximum of 30 is the one chosen. The described procedure is repeated for each considered time window.

### 3.2. Indicators of group dynamics

Four measures are considered to analyze group conformation dynamics; MST total distance, MST average path and number of identified groups, and the number of times two countries where in the same group.

Total distance (Equation 3) corresponds to the total sum of the MST links with the lowest connection cost (taking into account their respective weights). When comparing two MSTs, the one with the lower sum of links has a lower connection cost, reflecting a major coincidence in countries disease evolution.

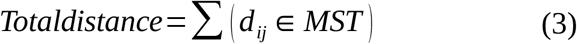

In the case of analysis, average path refers to the average distance in the evolution of the disease between two countries of the considered MST.

A lower average geodesic indicates that on average, any two countries of the network had more similar evolution in COVID – 19 contagions. The geodesic between nodes i and j is the shortest path between them (Jackson, 2010). Equation 4 reflects how to calculate average distance, for which it is necessary to define geodesics concept. From all possible paths between i and j in which any node does not figure more than one time, the geodesic is the shortest. When considering the I geodesics of the MST, the i-th one is represented as *n*_*i* ∈ (1, *I*)_.

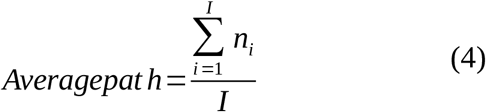

The number of identified clusters in each time window is determined as the highest number of suggested groups according to pseudo T^2^and CH cutting criteria. A smaller number of groups implies the presence of countries where the disease evolves in a similar way but that differs from how it evolves in other groups. The increase in the number of groups implies difficulties in identifying groups of countries that differ from others according to the evolution of the disease.

Finally, the number of times two countries are in the same group can provide information on groups’ stability. If the identified groups are constant (two nodes are always – 95 times - or never – 0 times – in the same group), the similarity in disease evolution between countries does not change in time, and otherwise, it is dynamic.

### 3.3. Communities identification

Subsection 3.1 procedure is repeated in every time window, obtaining information about which countries belong to the same group. Intensity in similarity of disease evolution is estimated by the number of times two countries were in the same group. Given that 95 time windows of 150 days amplitude that move one day at a time are considered, the lowest possible intensity between two countries is 0 and the highest 95. With this information, an adjacency matrix and the corresponding network is constructed.

Community identification is done by Louvain algorithm (Blondel et al., 2008). This method aims to achieve the maximum modularity (Equation 5), that represents the difference between observed links and the expected ones by the assumption that communities structure is independent from links formation over total links. Modularity takes values between −1 and 1, with 0 meaning that communities are independent of the amount of internal links, 1 that links are only formed inside the community and −1 that links are outside the community.

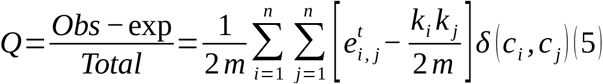

where *n*is the number of nodes in the network (124), 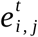 the element *i, j* of the adjacency matrix, 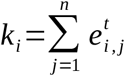 the sum of edges related to node *i*, 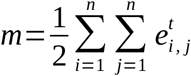 the community of node *i* and *δ* (*c*_*i*_, *c* _*j*_) the Kronecker delta that takes value 1 if *c*_*i*_=*c*_*j*_ and 0 if not.

The procedure starts with n communities of one node each, which is moved to neighboring communities if it implies an increase of modularity. First, community structure is obtained when these movements do not produce a major modularity. Then the procedure is repeated considering the formed communities as nodes and the sum of edges as links. Algorithm ends when the movement of nodes between communities does not increase modularity.

### 3.4. Communities characterization

Communities are characterized in several aspects. In particular, socioeconomic, demographic and geographical indicators are considered. Other information related to the pandemic, such as number of deaths and taken actions are also incorporated (more specific information is provided in Appendix 1).

## 4. Results

This section presents the results in 3 subsections. The dynamics of the groups identified in each time span is analyzed in the first subsection. The second subsection presents the main observations of the intensity network in similarity of disease evolution and the identified communities. Finally, the third section presents the results’ robustness.

### 4.1. Evolution of groups in time windows

Figure 1 shows the evolution of the number of identified clusters in the 95-time windows considered (purple) and the evolution of the MST total distance (orange). A wide variation in the number of groups is observed; note that the time windows move one day at a time, the maximum number of groups in each time window is 30 and the lowest number of identified groups is 3. In particular, there seems to be a cyclical behavior around a decreasing trend, with increases in the number of groups every 20 days approximately (corresponding to windows 21, 43, 58, 74 and 95). Total network distance increases in the first 20 time windows and decreases after. This means that seen as a whole, the trajectories of COVID-19 in the countries considered began to approach from the time window 20. Additionally, Figure 2 shows the evolution of networks average path (red) in comparison with the evolution of identified groups (purple). The formation of a smaller number of groups seems to be related to a major average path. To sum up, the identification of a smaller number of groups is related to moments in which the evolution of the disease was more similar between some countries and widened among others. Moreover, this reflects that evolution of the COVID–19 has been highly dynamic, changes in the interaction were fast, reflecting a high diversity of interacting trajectories that move closer to and away from each other and highlighting the importance to analyze it as a complex system.

**Figure 1:**
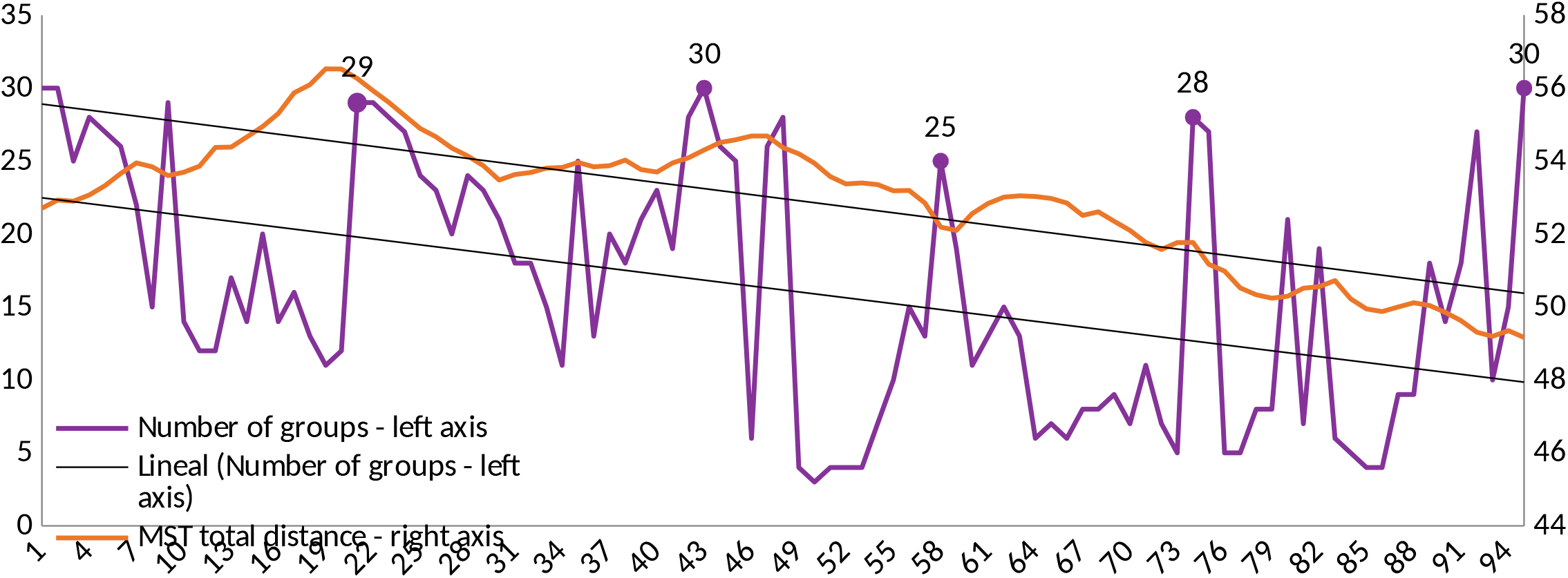
Number of identified groups and MST total distance in each time window Source: own construction based on OWID data.

**Figure 2:**
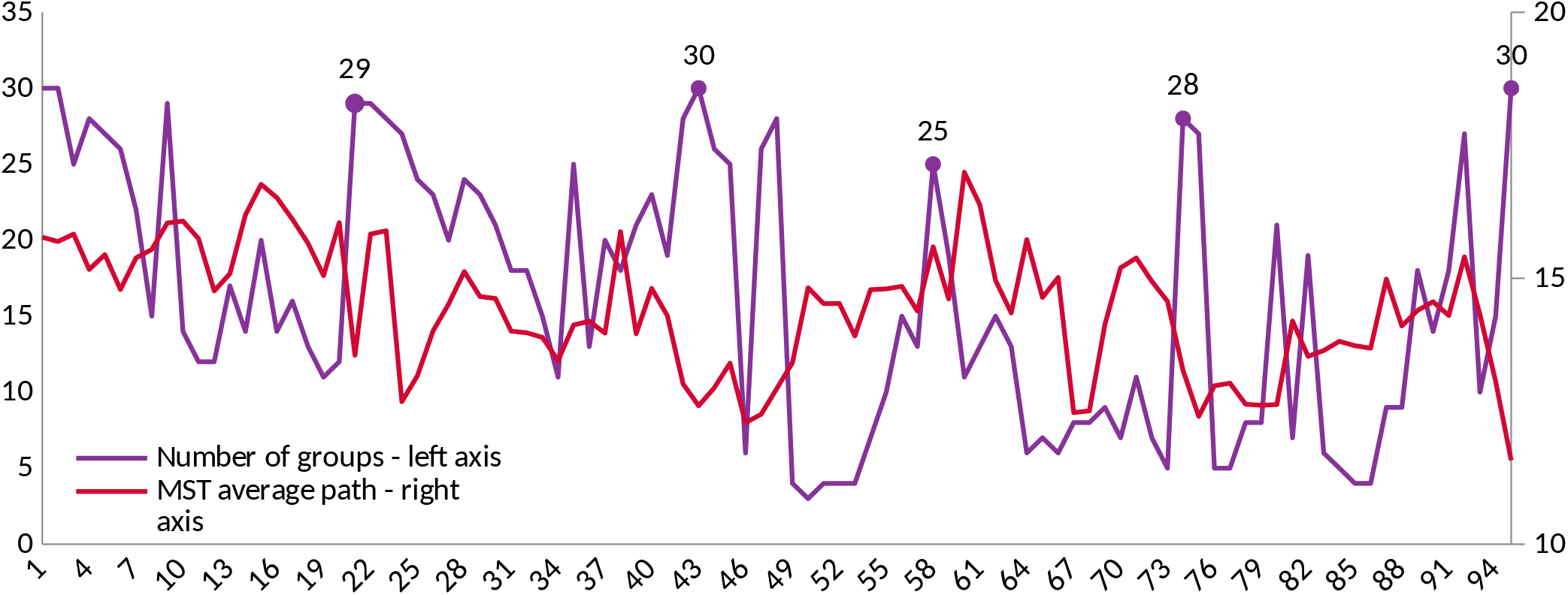
Number of identified groups and MST average path in each time window Source: own construction based on OWID data.

Likewise, the formation of groups of similarity in the evolution of the disease is also dynamic, since only in 5.17% of the cases the groups were static (see Figure 3). Note that most cases corresponds to a situation in which two countries are never in the same group (389 in 7626, 5,1% of the cases), while it is rare to find countries that belong to the same group in all considered time windows (5 in 7626, 0,07% of the cases).

**Figure 3:**
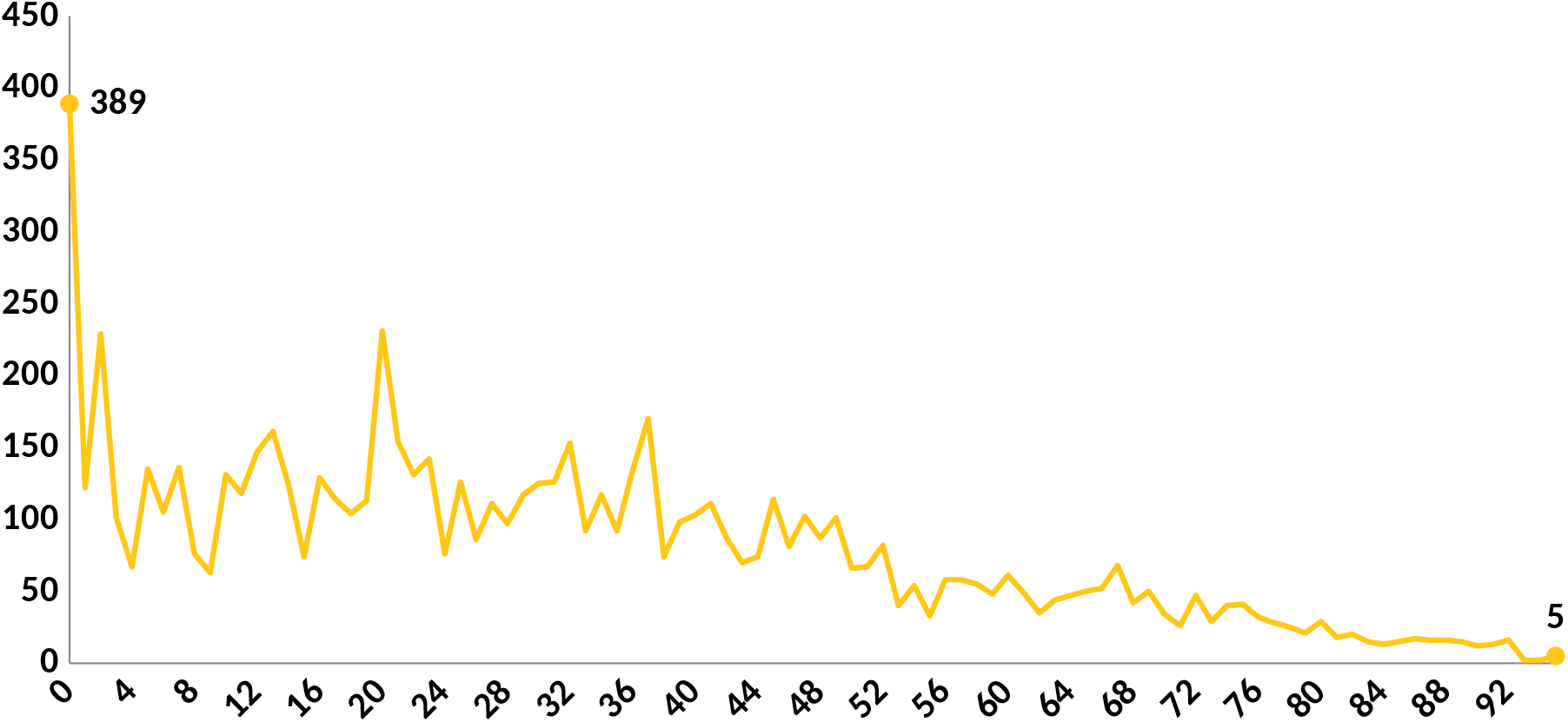
Number of times two countries where in the same group per time window Source: own construction based on OWID data.

### 4.2. Communities of countries by the intensity of similarity in the disease evolution

Three communities of countries were identified from the intensity of similarity in the disease evolution network (see Appendix 2), represented in Figure 4. Community 1 is conformed by 56 countries, community 2 of 59 and community 3 has only 9 members.

**Figure 4.**
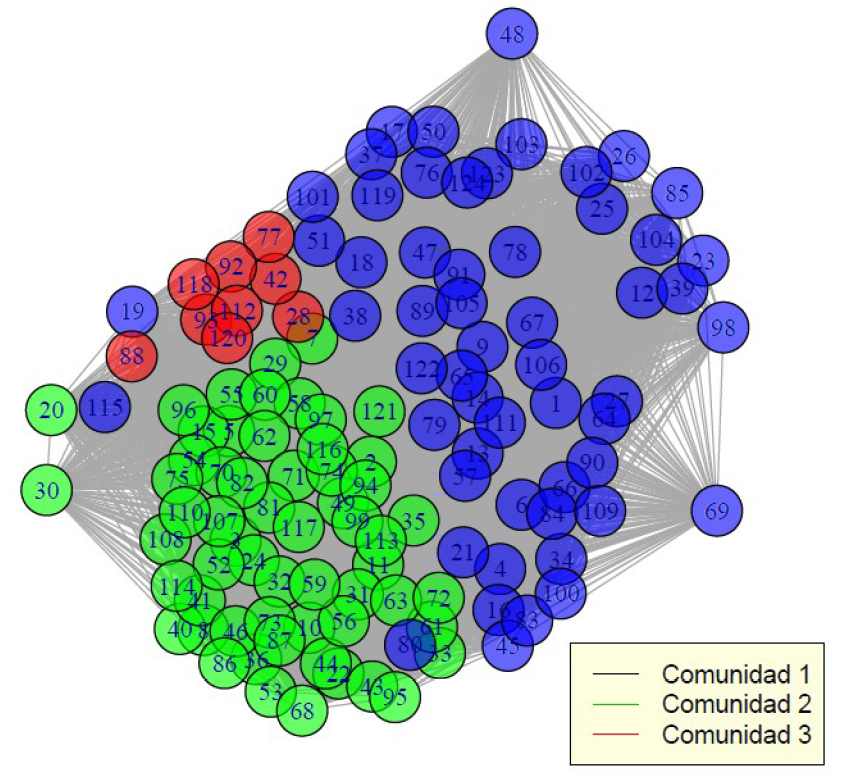
Intensity of similarity in the disease evolution network Source: own construction based on OWID data. Appendix 2 presents countries corresponding to each number.

As Figure 5 shows, the evolution of COVID–19 rolling 7-day average of daily new cases per million people in most of the community 1 was more constant than in the other communities. Even when individual trajectories were different between members (some had one wave, others two or more and levels are wide), the disease in these countries seems to oscillate in a smaller range in comparison to countries from the other communities. Average group trajectory presents a low positive trend. Initial spread of the disease seems to have been less abrupt than in other communities, with a more gradual increase in the number of cases in several countries. Community 2 is composed by countries in which the new confirmed cases per million had at least one phase of wide variation. Some of these countries had an important and fast increase in the number of daily cases at the beginning of infections, in others it happens during the middle of the considered time span. It is also possible to identify some countries with a slower growth in the number of cases, but with an important difference between days with small and high number of daily contagions. Inmost of the community members, at least one pronounced wave can be seen, but for some the number of daily new cases stabilizes at a high level after surge. On average, the community presents a first high and short wave of contagions, followed by a positive trend (higher than in community 1).

**Figure 5.**
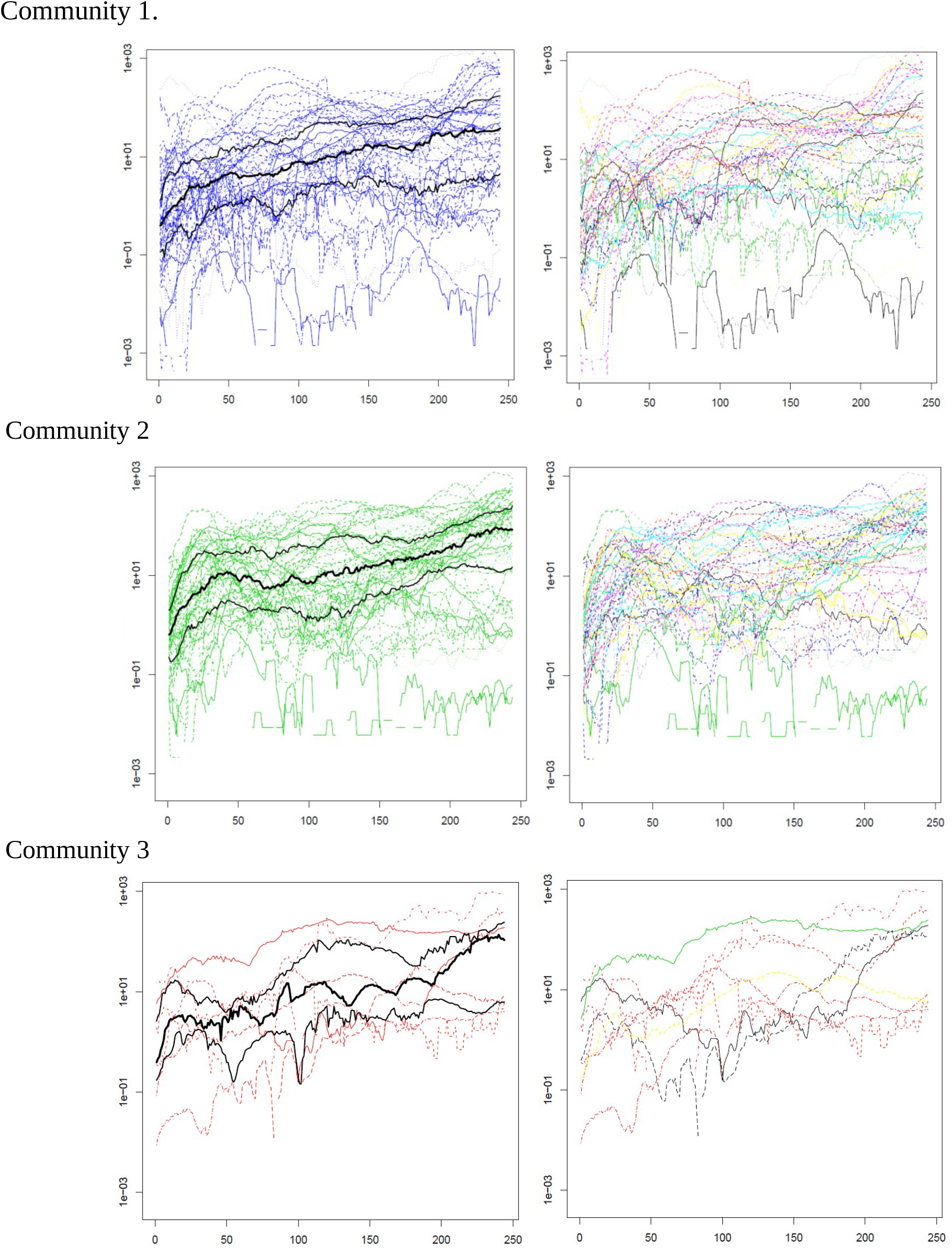
Rolling 7-day average of daily new confirmed COVID-19 cases per million people by country and community. Source: own construction based on OWID data. Black central lines correspond to group median; high and low black lines represent the upper and lower quartiles respectively.

Average disease evolution of community 3 is the most unstable, and fluctuates around a positive trend. The first wave of contagions seems to have been less pronounced and shorter than in community 2 for most members, additionally, longer phases of increase in the number of daily cases seems to be presented. Given that average evolution presents a positive trend in all communities, the number of daily cases has not yet entered a phase of decline. These results suggest that the number of infections will continue for a while.

Despite having similarities in relation to contagion dynamics, the identified communities are different in several aspects. Considered in average, each community has very different characteristics in terms of demographic, socioeconomic and geographical indicators, actions taken in relation to COVID–19, and in the impact of the disease (measured by the number of total deaths per million of inhabitants by December 1^st^).

Table 1 presents communities demographic and socioeconomic information that is considered as relevant in relation to the disease evolution. In all considered measures, communities have countries that belong to at least 3 quartiles of the variable distribution along all considered countries. This indicates that, even when there are characteristics in which some quartile have more countries, communities are diverse in all considered aspects.

**Table 1.** Communities’ demographic and socioeconomic data

|  |  | community 1 | community 2 | community 3 |
| --- | --- | --- | --- | --- |
| Average population density |  | 326 | 182 | 96 |
| Number of countries per quartile of population density | 1 | 14 | 15 | 1 |
|  | 2 | 10 | 16 | 5 |
|  | 3 | 13 | 16 | 2 |
|  | 4 | 19 | 11 | 1 |
| Median age |  | 33 | 35 | 29 |
| Number of countries per quartile of median age | 1 | 16 | 11 | 3 |
|  | 2 | 13 | 14 | 3 |
|  | 3 | 10 | 19 | 1 |
|  | 4 | 14 | 15 | 1 |
| Average GDP per capita* |  | 20,545 | 26,327 | 11,196 |
| Number of countries per quartile of GDP per capita | 1 | 16 | 9 | 4 |
|  | 2 | 15 | 13 | 2 |
|  | 3 | 12 | 16 | 3 |
|  | 4 | 11 | 19 | 0 |
| Average life expectancy |  | 75 | 76 | 72 |
| Number of countries per quartile of life expectancy | 1 | 14 | 12 | 5 |
|  | 2 | 15 | 15 | 1 |
|  | 3 | 14 | 14 | 3 |
|  | 4 | 13 | 18 | 0 |
| Average Human development index |  | 32.7 | 34.6 | 29.5 |
| Number of countries per quartile of Human development index | 1 | 17 | 11 | 3 |
|  | 2 | 16 | 12 | 2 |
|  | 3 | 9 | 18 | 3 |
|  | 4 | 14 | 17 | 0 |

Source: own construction based on OWID data. Note: Difference between sum of countries frequency and total countries per group at each variable are due to lack of data. \*Measure in constant 2011 international dollars, most recent year available.

In particular, community 1 has the highest average population density, with most countries in the 4^th^ quartile. In terms of median age, average per capita GDP, average life expectancy and average HDI, community 2 has the highest value. These communities present average values more similar to each other than to community 3 in most of the variables considered. Community 3 has the lowest population density and the youngest population (lowest average median age), but it is by far the most vulnerable in terms of the others variables.

In terms of geographical distribution there is not a clear pattern between communities. Most African and American countries belong to community 1 and there are more European countries in community 2. Even so, both communities have members from all continents. Members of community 3 are African, European and Asian countries, note that 99 of the 124 considered countries belong to this continents. All above considered, continent does not seem to be a crucial factor in determining communities of intensity in similarity of the evolution of COVID – 19 (see Table 2).

**Table 2.** Communities geographic data

|  | No. of countries | No. of countries by continent |  |  |  |  |  | No. of islands |
| --- | --- | --- | --- | --- | --- | --- | --- | --- |
|  |  | Africa | Asia | Europe | North America | Oceania | South America |  |
| Community 1 | 56 | 11 | 17 | 14 | 8 | 1 | 5 | 9 |
| Community 2 | 59 | 7 | 18 | 24 | 5 | 1 | 4 | 10 |
| Community 3 | 9 | 3 | 2 | 3 | 1 | 0 | 0 | 0 |
| Total | 124 | 21 | 37 | 41 | 14 | 2 | 9 | 19 |

Source: own construction based on OWID data.

Additionally, figure 6 shows how communities locate in the globe. As it can be seen, communities are spread in a wide range of latitudes and their members are not necessarily border.

**Figure 6.**
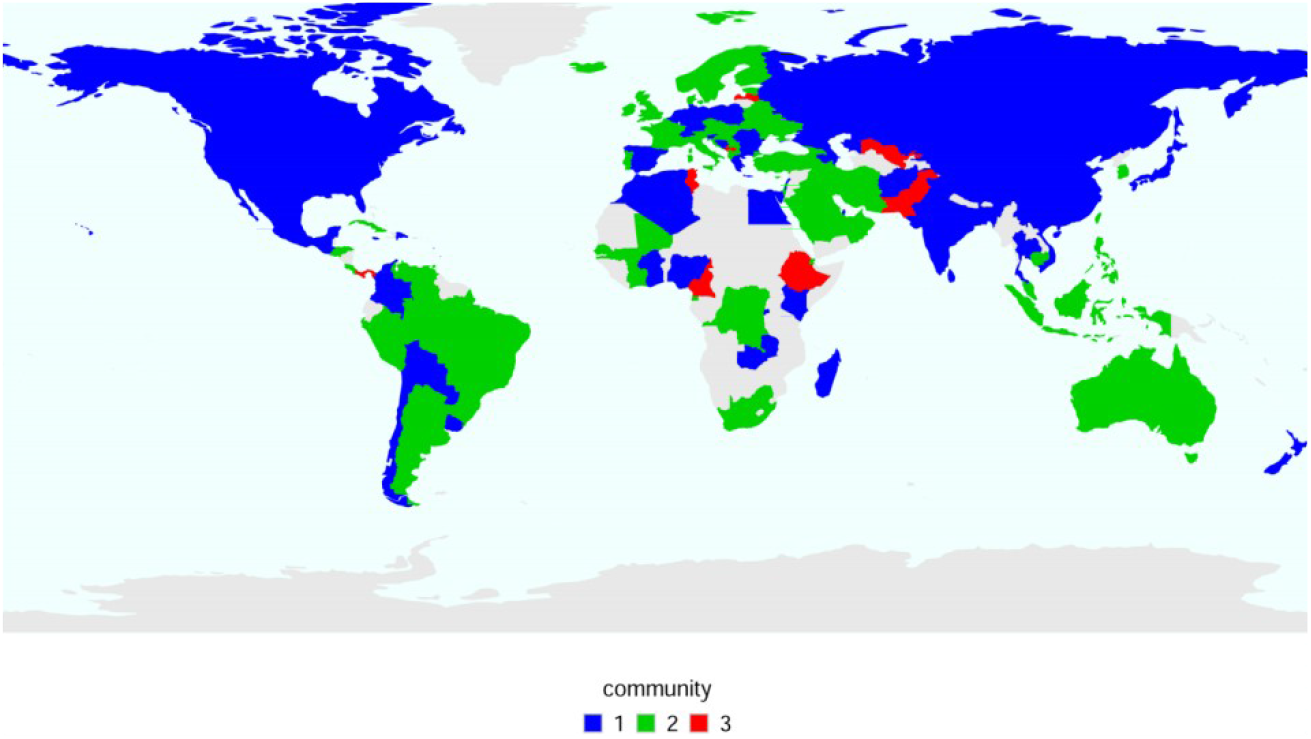
Communities map Source: own construction based on OWID data.

Communities’ members are also diverse in terms of implemented actions to counteract the negative effects of the disease in order to control its propagation. Overall government responses were varied within each community, due to both containment and health measures as well as economic ones (see table 3).

**Table 3.**
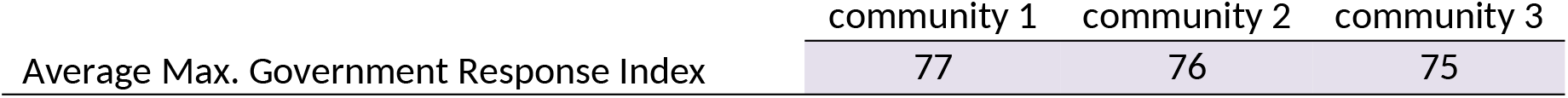
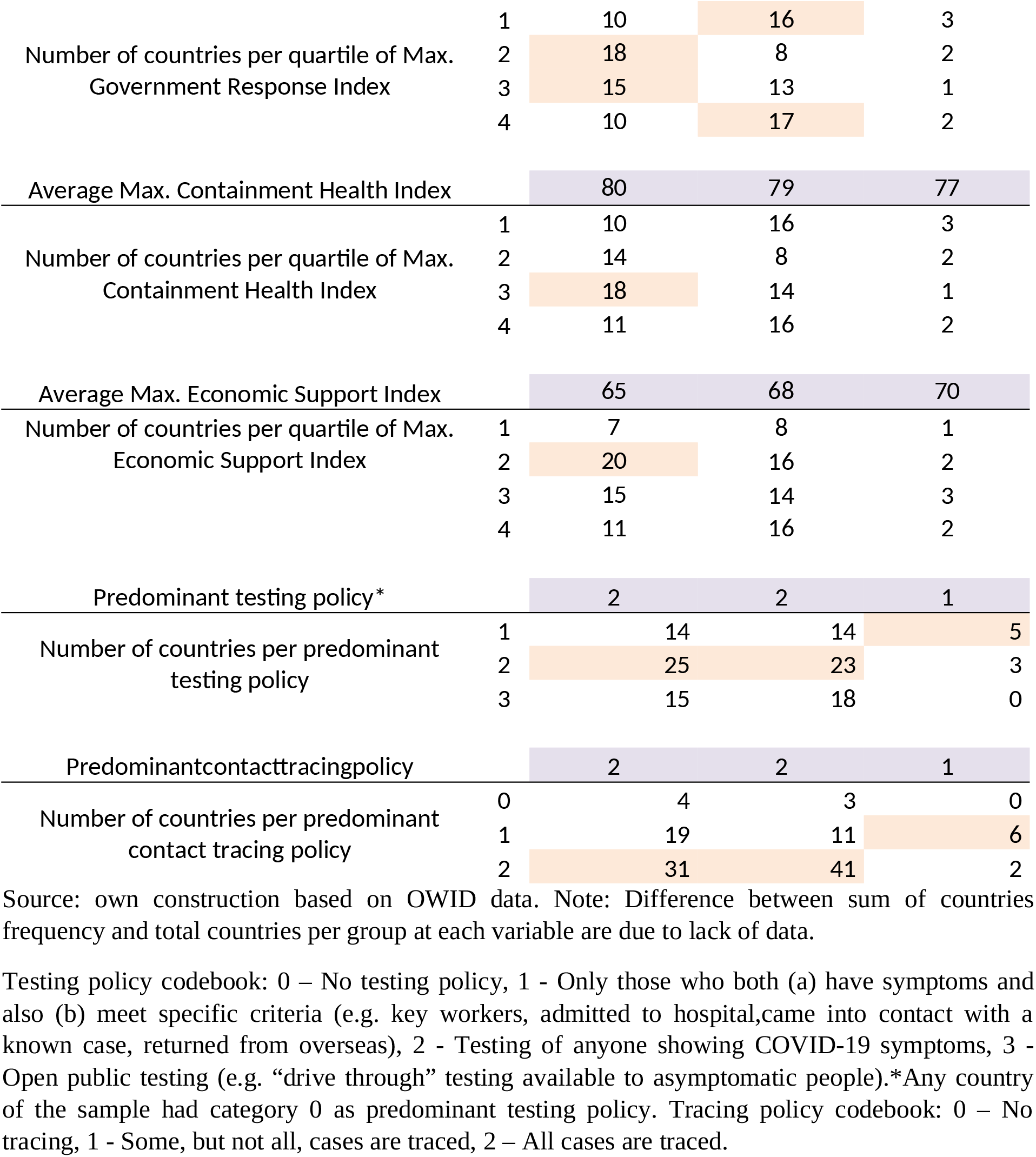
Countries actions in relation to COVID – 19

|  | community 1 | community 2 | community 3 |
| --- | --- | --- | --- |
| Average Max. Government Response Index | 77 | 76 | 75 |

|  |  |  |  |  |
| --- | --- | --- | --- | --- |
| Number of countries per quartile of Max. Government Response Index | 1 | 10 | 16 | 3 |
|  | 2 | 18 | 8 | 2 |
|  | 3 | 15 | 13 | 1 |
|  | 4 | 10 | 17 | 2 |
| Average Max. Containment Health Index |  | 80 | 79 | 77 |
| Number of countries per quartile of Max. Containment Health Index | 1 | 10 | 16 | 3 |
|  | 2 | 14 | 8 | 2 |
|  | 3 | 18 | 14 | 1 |
|  | 4 | 11 | 16 | 2 |
| Average Max. Economic Support Index |  | 65 | 68 | 70 |
| Number of countries per quartile of Max. Economic Support Index | 1 | 7 | 8 | 1 |
|  | 2 | 20 | 16 | 2 |
|  | 3 | 15 | 14 | 3 |
|  | 4 | 11 | 16 | 2 |
| Predominant testing policy* |  | 2 | 2 | 1 |
| Number of countries per predominant testing policy | 1 | 14 | 14 | 5 |
|  | 2 | 25 | 23 | 3 |
|  | 3 | 15 | 18 | 0 |
| Predominant contact tracing policy |  | 2 | 2 | 1 |
| Number of countries per predominant contact tracing policy | 0 | 4 | 3 | 0 |
|  | 1 | 19 | 11 | 6 |
|  | 2 | 31 | 41 | 2 |

Source: own construction based on OWID data. Note: Difference between sum of countries frequency and total countries per group at each variable are due to lack of data.
Testing policy codebook: 0 – No testing policy, 1 - Only those who both (a) have symptoms and also (b) meet specific criteria (e.g. key workers, admitted to hospital, came into contact with a known case, returned from overseas), 2 - Testing of anyone showing COVID-19 symptoms, 3 - Open public testing (e.g. “drive through” testing available to asymptomatic people). \*Any country of the sample had category 0 as predominant testing policy. Tracing policy codebook: 0 – No tracing, 1 - Some, but not all, cases are traced, 2 – All cases are traced.

More differences can be identified in terms of predominant testing and contact tracing policies (see table 3). In communities 1 and 2, the most frequent testing policy between January 22 and December 1^st^ was to test anyone showing symptoms, while in community 3 people had to also meet specific criteria. Tracing policy has also been more rigorous in communities 1 and 2; the most frequent tracing policy between January 22 and December 1^st^ was to traced almost all cases, in community 3 it was to trace some cases. Given that in average, communities 1 and 2 had smoother evolution of the disease than community 3, testing and tracing policies seems to be important in controlling contagions. Even so, communities are not homogeneous in the implementation of these policies, indicating that these measures do not guarantee the control the COVID–19 spread.

Having a similar evolution of the disease did not imply different results in terms of deaths, since average community registered deaths at December 1^st^ per million are similar between communities. Moreover, higher averages on healthcare access and quality index were not reflect in lower deaths averages (see table 4).

**Table 4.**
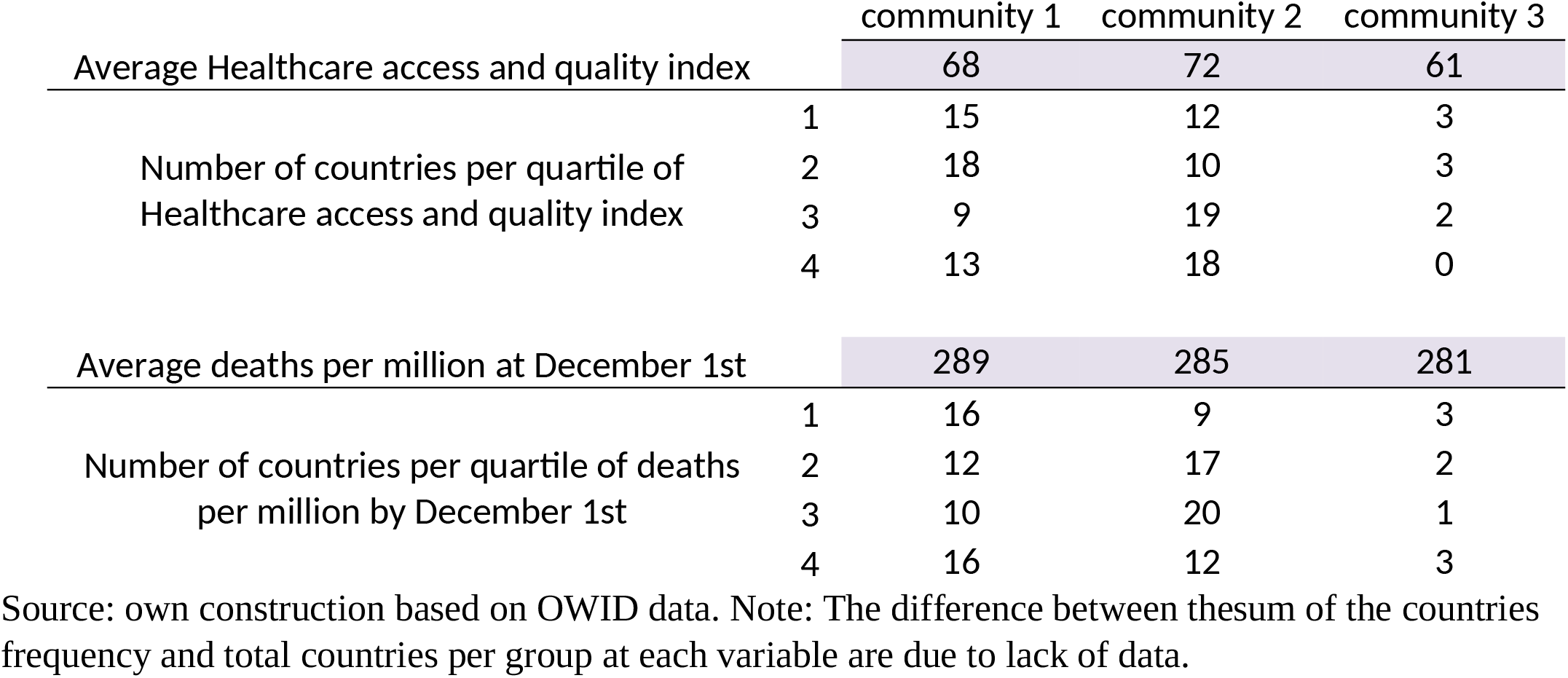
Healthcare system and deaths per million.

### 4.3. Results Robustness

Variations in the maximum number of groups allowed in each time window can have effects in formed communities. In this study, results were obtained for a maximum of 10, 20 and 30 groups per time window. Identified communities were stable; only 10 out of 124 countries belonged to a different community in at least one of the exercises performed. Table 5 presents observed countries community changes when varying the maximum of allowed groups per window. Appendix 3 presents countries’ values and quartiles per considered variable, confirming that communities are still diverse when these changes in membership are considered.

**Table 5.** Country community changes by allowed group maximum in each time window.

|  | Max. No. of groups per time window |  |  |
| --- | --- | --- | --- |
|  | 30 | 20 | 10 |
| Belgium | 1 | 1 | 2 |
| Bolivia | 1 | 3 | 1 |
| Bangladesh | 1 | 2 | 2 |
| Andorra | 1 | 3 | 1 |
| Chile | 1 | 1 | 2 |
| Morocco | 1 | 1 | 2 |
| Togo | 1 | 3 | 3 |
| Australia | 2 | 3 | 3 |
| South Africa | 2 | 3 | 3 |
| Montene-<br>gro | 3 | 2 | 3 |

Source: own construction based on OWID data.

## 5. Conclusions

Measuring the intensity of similarity in the disease evolution, three communities of countries were found, with distinctive aspects in their average behaviors. Community 1 presents the most controlled path, while in Community 3 average contagions were more unstable. Community 2 presents an intermediate average evolution, with an initial wave followed by a more stable path. Even so, communities’ members were diverse in several aspects, and the three communities present particular cases that do not follow the average behavior. All communities presented positive trends, which suggests that the number of infections will continue for a while. Furthermore, the multiplicity of dynamics suggests that the impact on the national economies will be heterogeneous, with some countries less affected. This might be observed in the levels of unemployment. In this regard, Milani (2020) finds that the behavior of unemployment across countries has been very heterogeneous. In Latin American countries, and even in the US, socially disadvantaged populations cannot follow the order to stay at home. Thus, the issue of income inequality turns to be the main obstacle for a complete and effective implementation of social distancing measures. Even so, each community is integrated by countries from different GDP per capita levels.

Our results confirm the initial assumption. The spread of COVID-19 is a complex phenomenon, characterized by both a multiplicity of patterns and non-linear dynamics. This point suggests, on one hand, the presence of diverse drivers behind the spread of the COVID-19 and that public policy responses should not be homogeneous, but should rather be adapted to different contexts. However, it is important to note that communities with a more stable evolution were composed by a majority of countries implementing stricter testing and contact tracing policies.

Given the complexity of the phenomenon, we consider relevant to distinguish between short-term and medium-term interventions. The implementation of lockdowns is an example of the former. When discussing the effect of lockdowns, the available literature seems to show contradictory results. However, since COVID propagation is a complex phenomenon, we consider that there is no such contradiction, but that the results should be analyzed within their specific contexts. Note that countries from the same community implemented different levels of maximum containment health policies.

In particular, policy makers should specify in their objective function which segments of the population are being targeted and which indicators are been used. In this regard, as Ehlert (2020) points out, policymakers should consider the identification of risk groups when designing their response strategies. In addition, Huang et al. (2020) recommend policy makers to guarantee that the impacts of public policy include the interests of the socially disadvantaged groups. It is also important to consider the existence of trade-offs within the objective function, since minimizing both COVID-19 propagation and economic impacts might be two conflicting goals. On this line, Mahasinghe et al. (2020) suggest that the lockdown strategies are non-linear. For example, in the case of Italy, Ciminelli and Garcia-Mandio (2020) offer evidence that the shutdown of service activities is effective in reducing COVID-19 mortality, although shutting down factories is less effective. Thus, they recommend closing down services, however the government should be more careful when considering closing down factories, particularly given the social costs derived from halting production. In the case of Chile, Asahi et al (2020) found out that localized lockdowns were associated to up to 15% drop in the local economic activity.

Regarding the service sector, it would be important to have a second thought when considering a complete shut down in the tourism sector, as Ehlert (2020) suggests. In his study of Germany’s 401 counties, he finds no evidence of the tourism sector as a driver of infection or death rates. At this point, it would be important to consider that tourism encompasses a wide range of activities and that many tourists travel in order to isolate themselves during their leisure time. Thus, this activity might be favoring conditions for social distancing. In the case of Latin American countries, in which the rate of infections and mortality has been growing, policy makers might stimulate self-isolation through promoting the use of resorts for risk groups.

The best options to counteract the harmful effects of a pandemic may be having strong health systems with contingent capacity to deal with unforeseen events or available resources capable of a rapid expansion of its capacity. Regarding the medium-term interventions, the expansion of health services should be considered. In Latin American countries, an important proportion of population has diabetes and hypertension, which increase fatality rates. The policy recommendation is a robust public-funded health system with wider accessibility, as Sherpa (2020) and Gandjour (2020) suggest. There should be an increase in public health funding, as well as in the number of doctors per population and bed availability. Alternatively, countries could have available resources with the capacity to rapidly strengthen the health system. Ciminelli and Garcia-Mandio (2020) point out that a robust health care system is the only way to prevent a huge death toll. Their results for Italy show that locations at 10 km from the closest intensive care unit had up to 50% higher mortality.Even when communities are heterogeneous in terms of total deaths and healthcare access, lack of information on the number of cases that required hospitalization in several considered countries limits our results in this aspect. Additionally, policy makers should pay attention to bottlenecks in the access to health services, since it has been observed that isolating health areas just for COVID-19 patients has complicated the treatment of other health problems. In order to avoid additional disruptions in health treatments, Degeling et al (2020) recommend the restoring of health service to normal levels as soon as possible. Another relevant medium-term intervention would be improving the health of the population via the intake of more fruits and vegetables. Since it implies a drastic change in dietary habits, it might require time and an important effort by governments and regulatory agencies. This intervention might be particularly relevant in Latin American countries, in which obesity and related co-morbidities are common. In a study of the population in the Nordic countries, Mol and Karnon (2020) found the that the existence of low obesity rates facilitates a more liberal social distancing policy and that the impact of consumption of fruits and vegetable on deaths rates would be comparable or even better than strict lockdowns. The study shows evidence that as the disease progresses, the pattern diversifies. As there are no clear general characteristics that allow the different trajectories to be identified, the best response in the medium term is a well-prepared and strong health system.

## Data Availability

The data that support the findings of this study are openly available in Our World in Data.

https://ourworldindata.org/

## Acknowledgments

Earlier versions of this article, to which the authors have contributed equally, were presented at the 5th - Webinar COVID-19 Forecast and Prediction - September 18th-19th, 2020 (https://www.covid-19-response-webinar.org/index.php?title=5th_Webinar_-_Forecast_and_Prediction) organized by the Karolinska Institutet, Stockholm, Sweden.

The authors wish to thank the participants of the GIDE seminar and the colloquium of the Lateinamerika-Institut (LAI) - Freie Universität Berlin for providing helpful and constructive comments on earlier versions of this article

## Appendix 1. Complementary variables codebook

|  | Description - Data source |
| --- | --- |
| Populationdensity | Number of people divided by land area, measured in square kilometers, most recent year available - World Bank World Development Indicators, sourced from Food and Agriculture Organization and World Bank estimates |
| Median age | Median age of the population, UN projection for 2020 - UN Population Division, World Population Prospects, 2017 Revision |
| GDP per capita | Gross domestic product at purchasing power parity (constant 2011 international dollars), most recent year available - World Bank World Development Indicators, source from World Bank, International Comparison Program database |
| Lifexpectancy | Life expectancy at birth in 2019 - James C. Riley, Clio Infra, United Nations Population Division |
| HDI | Summary measure of average achievement in key dimensions of human development: a long and healthy life, being knowledgeable and have a decent standard of living - United Nations Development Programme (UNDP) |
| Max Gobermentrespon-<br>ceindex | The Oxford Covid-19 Government Response Tracker tracks individual policy measures across 18 indicators. Highest daily value is taken. - Thomas Hale, Sam Webster, Anna Petherick, Toby Phillips, and Beatriz Kira (2020). Oxford COVID-19 Government Response Tracker, Blavatnik School of Government. |
| Max conteintment and<br>health index | This is a composite measure based on eleven policy response indicators including school closures, workplace closures, travel bans, testing policy and contact tracing, rescaled to a value from 0 to 100 (100 = strictest). If policies vary at the subnational level, the index is shown as the response level of the strictest sub-region. Highest daily value is taken. - Thomas Hale, Sam Webster, Anna Petherick, Toby Phillips, and Beatriz Kira (2020). Oxford COVID-19 Government Response Tracker, Blavatnik School of Government. |
| Max economicindex | The Oxford Covid-19 Government Response Tracker tracks individual policy measures across 2 indicators, income support and debt/contract relief. Highest daily value is taken. - Thomas Hale, Sam Webster, Anna Petherick, Toby Phillips, and Beatriz Kira (2020). Oxford COVID-19 Government Response Tracker, Blavatnik School of Government. |
| Testingpolicy | Applied criteria to carry out COVID-19 tests - Thomas Hale, Sam Webster, Anna Petherick, Toby Phillips, and Beatriz Kira (2020). Oxford COVID-19 Government Response Tracker, Blavatnik School of Government. |
| Contacttracing | Applied criteria to carry out COVID-19 contact tracing - Thomas Hale, Sam Webster, Anna Petherick, Toby Phillips, and Beatriz Kira (2020). Oxford COVID-19 Government Response Tracker, Blavatnik School of Government. |
| Acceesshelth and quality | The Healthcare Access and Quality (HAQ) Index is measured on a scale from 0 (worst) to 100 (best) based on death rates from 32 causes of death that could be avoided by timely and effective medical care (also known as 'amenable mortality'). - Global Burden of Disease Study 2015. Global Burden of Disease Study 2015 (GBD 2015) |
| Total deaths per million<br>at Dec. 1st | Total deaths attributed to COVID-19 per 1,000,000 people by december 1st - COVID-19 Data Repository by the Center for Systems Science and Engineering (CSSE) at Johns Hopkins University |

Source: own construction based on OWID and GitHub data. December 8 search in: https://github.com/owid/covid-19-data/blob/master/public/data/owid-covid-codebook.csv, https://ourworldindata.org/coronavirus-testing, https://ourworldindata.org/grapher/healthcare-access-and-quality-index?time=2015, https://github.com/OxCGRT/covid-policy-tracker/blob/master/documentation/index_methodology.md and https://github.com/OxCGRT/covid-policy-tracker/blob/master/documentation/codebook.md

## Appendix 2 Identified communities

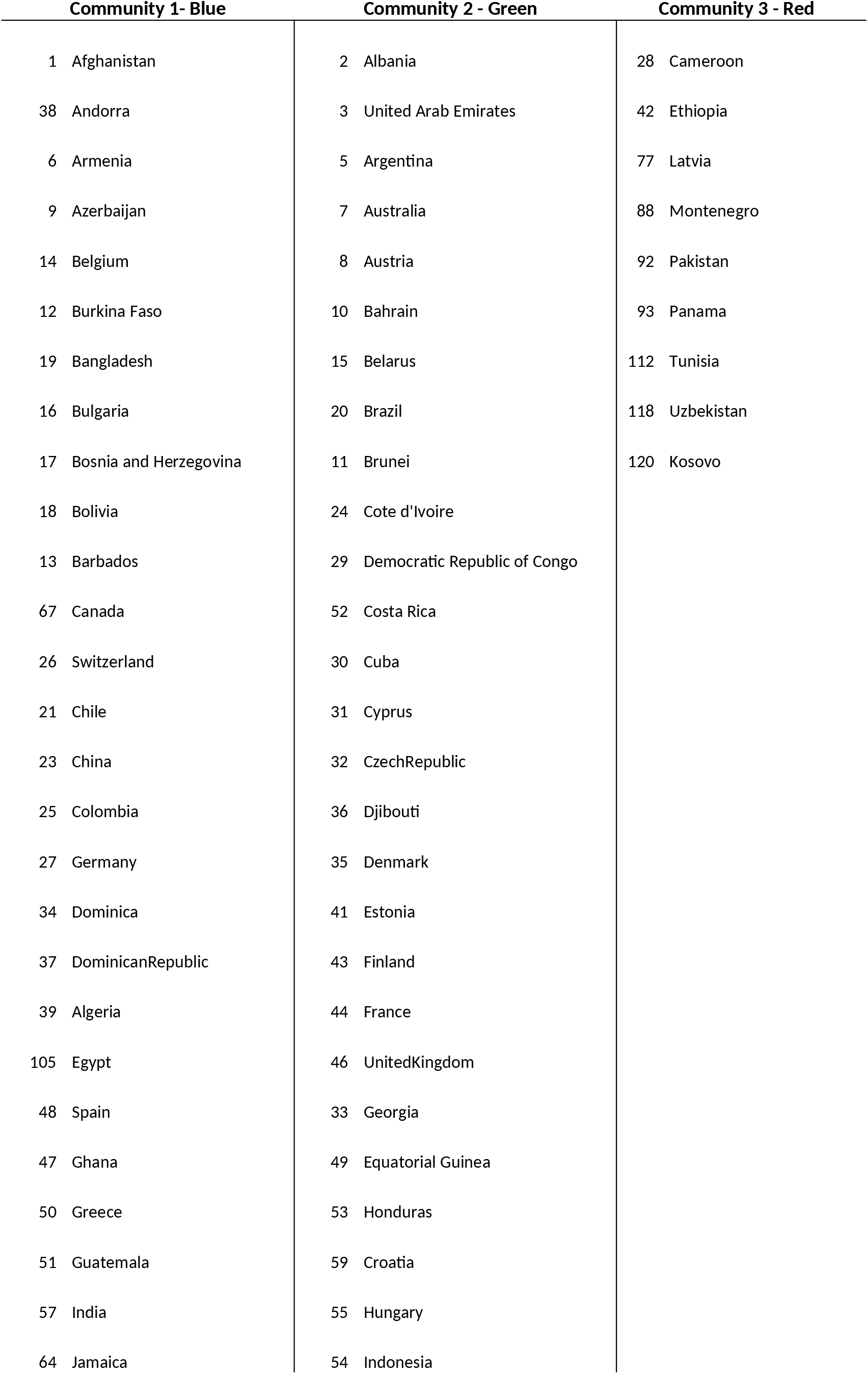
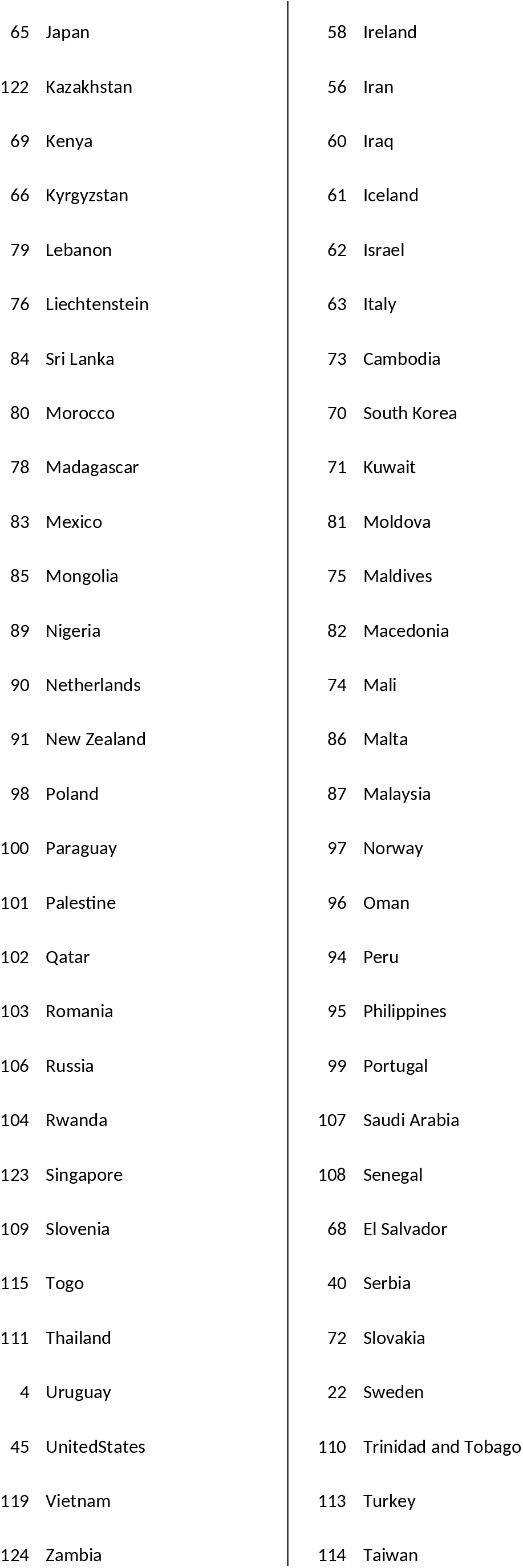
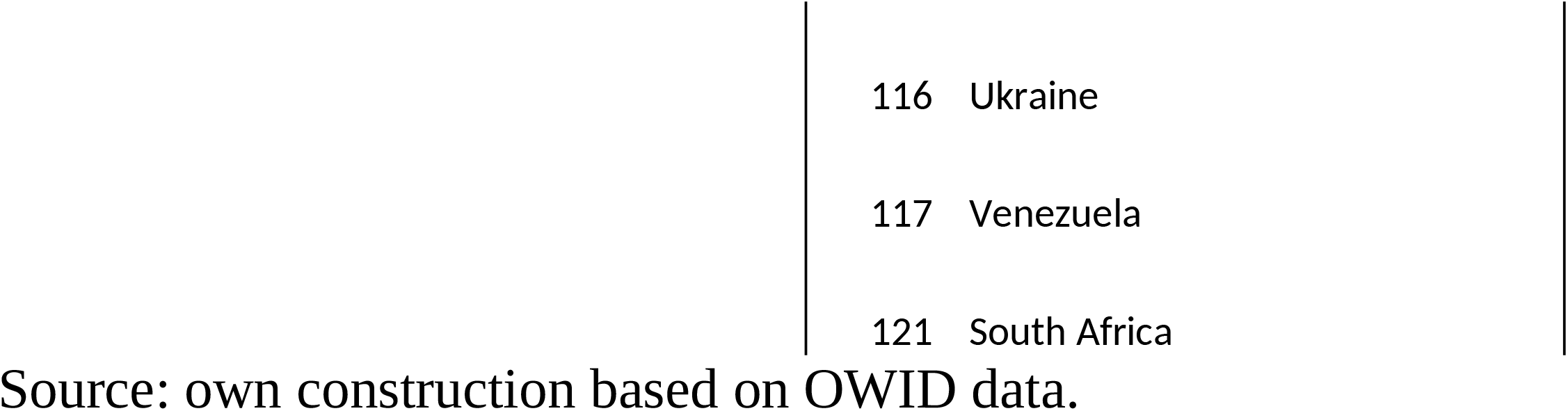

## Appendix 3: Countries data

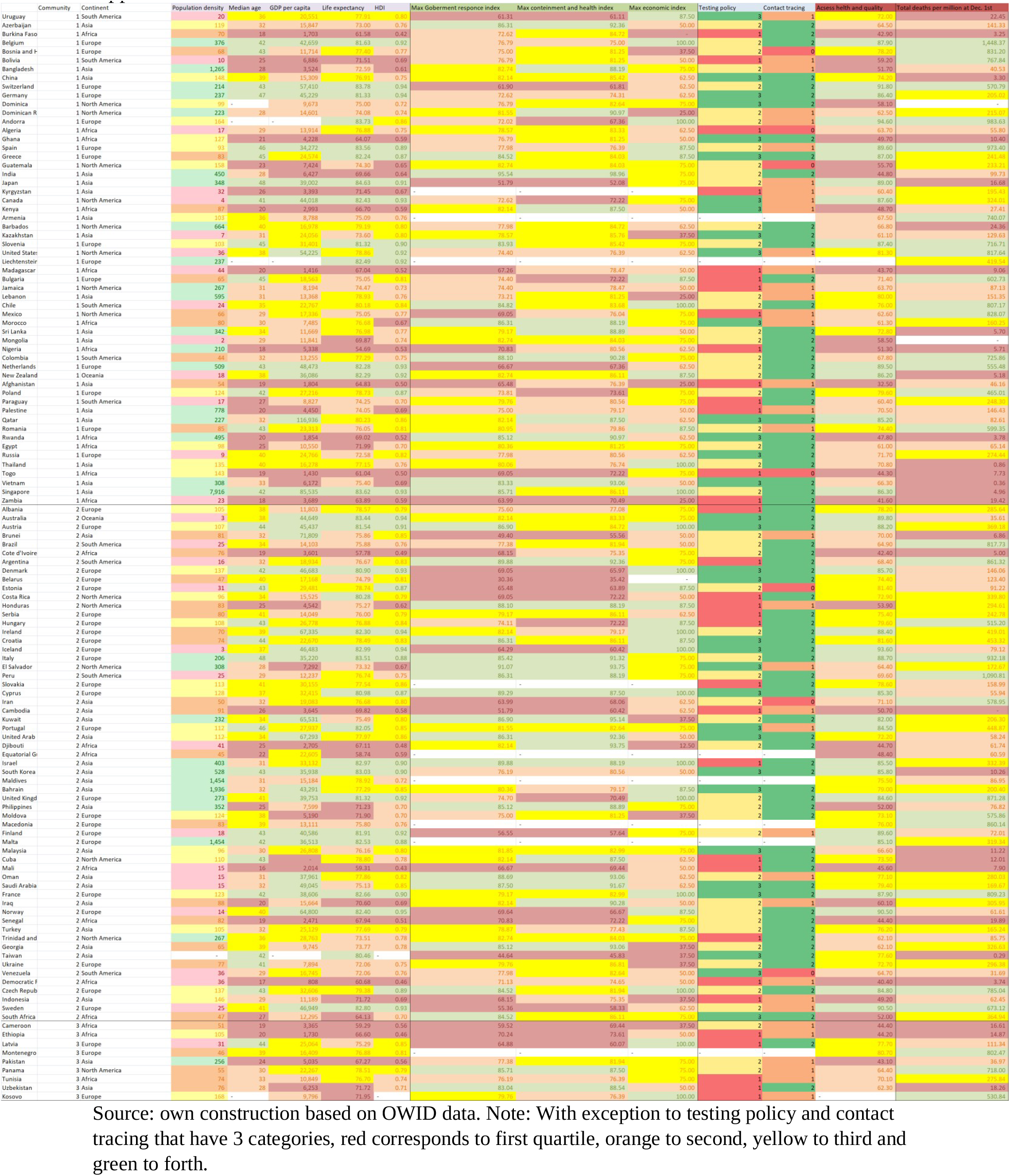

